# Post-COVID-19 Functional Status: Relation to age, smoking, hospitalization and comorbidities

**DOI:** 10.1101/2020.08.26.20182618

**Authors:** Aliae AR Mohamed Hussein, Islam Galal, Mahmoud M Saad, Hossam Eldeen E Zayan, Mustafa Z Abdelsayed, Mohamed M Moustafa, Abdel Rahman Ezzat, Radwa ED Helmy, Howaida K Abd Elaal, Karim Aly, Shaimaa S Abdelrheem

## Abstract

**Rational:** Recently, a new “Post-COVID-19 Functional Status (PCFS) scale” is recommended in the current COVID-19 pandemic. It is proposed that it could be used to display direct retrieval and the functional sequelae of COVID-19.

**Aim of the study:** To assess the Post COVID-19 functional status in Egypt and to evaluate if age, gender, comorbidities have any effect on functional limitations in recovered COVID-19 patients.

**Patients and methods:** A total of 444 registered confirmed COVID-19 patients were included. They were interviewed in our follow-up clinics or by calls and filled an Arabic translated PCFS scale in paper or online forms as well as their demographic and clinical data.

**Results:** 80% of COVID-19 recovered cases have diverse degrees of functional restrictions ranging from negligible (63.1%), slight (14.4%), moderate (2%) to severe (0.5%) based on PCFS. Furthermore, there was a substantial variance between the score of PCFS with age (P= 0.003), gender (P= 0.014), the duration since the onset of the symptoms of COVID-19 (P <0.001), need for oxygen supplementation (P<0.001), need for ICU admittance (P= 0.003), previous periodic influenza vaccination (P<0.001), smoking status (P < 0.001) and lastly the presence of any comorbid disorder (P <0.001).

**Conclusions:** Most of the COVID-19 recovered cases have diverse degrees of functional restrictions ranging from negligible to severe based on PCFS. These restrictions were affected by age, gender, periodic influenza vaccination, smoking, duration since symptoms onset, need for oxygen or ICU admittance, and lastly the presence of coexisting comorbidity.

## Introduction

Throughout history, there have been plenty of pandemics, however; the social response to COVID-19 is unparalleled. The world will certainly not be identical for a second time. It is assessed that almost four billion individuals are living in social segregation during this mother of all pandemics **(1)**.

Initially described in China in December 2019, a severe acute respiratory syndrome caused by coronavirus 2 (SARS-CoV-2) has spread all over the world. Most countries are still grappling and some are struggling with the problem, nowadays there was proved, 20.6 million confirmed cases of COVID-19, as well as 749K deaths worldwide (2). Egypt reported slightly over 95.963000 confirmed COVID-19 cases with 5085 deaths **(3)**. The new pandemic is injuring not only health organizations of several countries but also the financial prudence universal.

In the coming days, great stress will progressively comprise post-acute carefulness of those recovered cases from COVID-19. It is expected that COVID-19 may have a principal effect on the physical, mental, cognitive, and public health state, similarly in cases with minor disease exhibitions **(4)**. Preceding outbreaks of coronaviruses have been concomitant with persistent impairment in pulmonary function, muscle weakness, pain, lethargy, depressed mode, anxiety, vocational disorders, and impaired quality of life to various grades **(5,6,7)**.

It is fundamental to have a simple measure to monitor the progression of symptoms and the effect of these symptoms on the functional state of the affected patients. Because of the enormous number of COVID-19 recovered cases that necessitate strong follow-up, a simple and reproducible measure to categorize those patients complaining from sluggish or partial recovery would aid in guiding the deliberate use of medical funds and will also standardize research efforts. Recently, A group of investigators recommended an ordinal scale for evaluation of patient-relevant functional restrictions following an event of venous thrombo-embolism (VTE): the post-VTE functional status (PVFS) scale **(8, 9)**. It covers the full spectrum of functional consequences and focuses on both restrictions in usual activities and alterations in life-style in 6-scale scores. It is already known that there is a great frequency of pulmonary embolism, myocardial injury/ myocarditis and neurological dysfunctions, in severely ill cases with COVID-19 **(10, 11)**. That’s why **Klok and his colleagues** designed their novel “Post-COVID-19

Functional Status (PCFS) scale” (after slight adaptation) to be valuable in the existing COVID-19 pandemic **(12)**. The recommended new scale could be used upon hospital discharge, at 4-8 weeks after-discharge to display direct rescue, and at 6 months to evaluate functional residue. **Aim of this work is to:** Assess the Post COVID-19 functional status in Egypt by the PCFS scale and to evaluate if age, gender, comorbidities have any effect on functional limitations.

### Patients & methods

During the period from 15^th^ July to 13^th^ August 2020, patients were included if they had confirmed COVID-19 in the registry of Ministry of Health and Population in Egypt (positive or indeterminate COVID-19 PCR test, or presumed presence of Covid-19 based on clinical & radiological criteria). They were contacted and asked to fill online forms. Medical students, residents and volunteers evaluated patient’s symptoms and revised the submitted forms for missing data. We used an online format of Arabic translated Post COVID-19 functional scale. We recruited patients till the needed number of participant was achieved.

**Study design:** Cross-sectional study.

**Sample size:** was calculated using Epi info statistical package version 7. Based on, the following parameters for cross-sectional study; expected Post-COVID-19 cases 0.50, with acceptable margin of error 0.05, design effect 1, 95% confidence level. The required sample size will be 384 patients. It will be raised to 425 after considering 10% as a drop out (number of cases in Egypt on 18^th^ July 26, 2020=86474)

### The following data were collected

- The patient demographics and clinical data including age, gender, smoking status, **residence, seasonal influenza vaccination**, the presence of coexisting comorbidity quarantine status, **oxygen supplementation** and history of ICU admittance history were recorded
- **Post-COVID-19 Functional Status Scale (PCFS) scale (12)**: A questionnaire covering the entire range of functional limitations, including changes in life-style, sports, and social activities. If there was no limitation of activity, it was graded as grade 0, if there was a negligible effect on activities for patients was considered grade 1, whereas a lower intensity of the activities was considered as grade 2. Grade 3 accounted for the inability to perform certain accomplishments, forcing patients to structurally modify these. Finally, grade 4 was reserved for those patients with unembellished functional restrictions.
- **The study was approved by the ethical committee of Aswan Faculty of Medicine, Egypt and *registered in Clinicaltrial.gov: NCT04479293***.

### Statistical analysis

Data was coded and analyzed using the Statistical Package of Social Science software program, version ***26*** (IBM SPSS 26 Statistics for windows, Armonk, NY: IBM Corp). Data was presented as range, mean, standard deviation, for quantitative variables and frequency and percentage for qualitative variables. Comparison for qualitative variables was performed using Chi-square, while for quantitative variables, the comparison was conducted using. One-way ANOVA test, P values ≤ 0.05 was considered substantially significant.

## Results

### Demographic and Clinical features of the Study Sample

The study involved 444 participants. They were 192 males and 252 females, the mean age was 33.09±12.09 years, and the range was (18-86 years). Most cases (71.2%) resided in the urban areas versus (28.2%) lived in rural areas. 77.9% of participants were non-smokers, 13.1% were active smokers, while 9% were former smokers. 25% of cases had comorbid disorders. The mean duration since the onset of their symptoms was 35.31±18.75 days, 75.7% were admitted in hospitals, 20.7% required oxygen therapy, while 13.5% needed ICU as shown in ***Table (1)***.

**Table 1.**
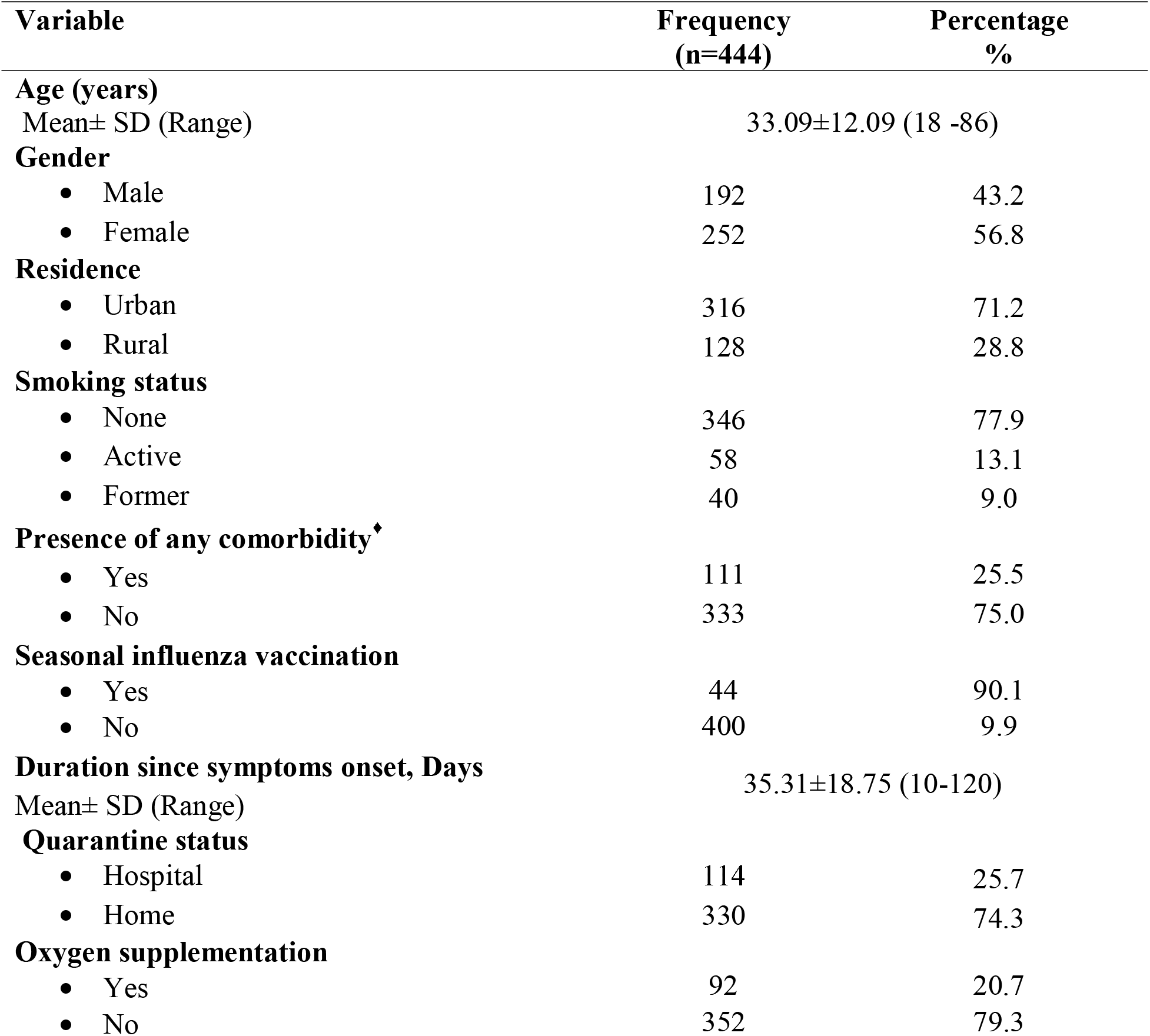
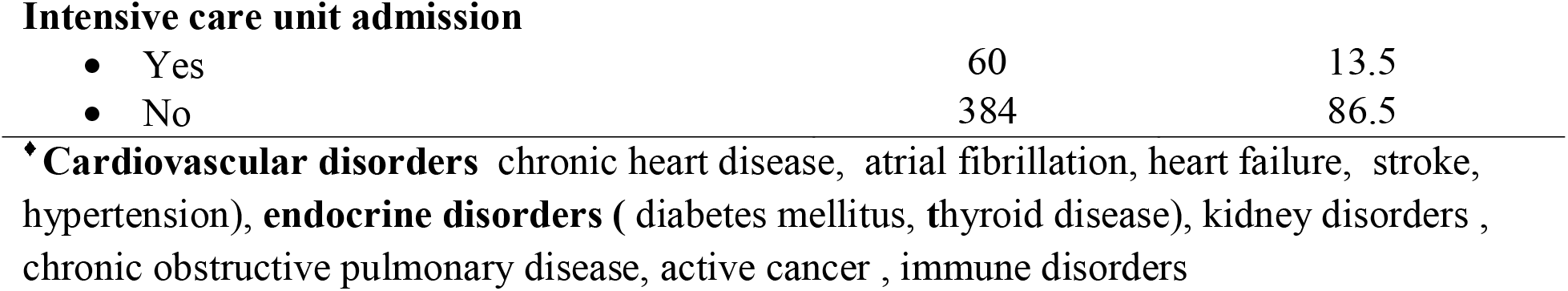
Demographic and Clinical Characteristics of COVID-19 recovered cases included in the study (n=444)

| **Variable** | **Frequency**<br>**(n=444)** | **Percentage**<br>**%** |
| --- | --- | --- |
| **Age (years)** |  |  |
| Mean± SD (Range) | 33.09±12.09 (18 -86) |  |
| **Gender** |  |  |
| • Male | 192 | 43.2 |
| • Female | 252 | 56.8 |
| **Residence** |  |  |
| • Urban | 316 | 71.2 |
| • Rural | 128 | 28.8 |
| **Smoking status** |  |  |
| • None | 346 | 77.9 |
| • Active | 58 | 13.1 |
| • Former | 40 | 9.0 |
| **Presence of any comorbidity<sup>♦</sup>** |  |  |
| • Yes | 111 | 25.5 |
| • No | 333 | 75.0 |
| **Seasonal influenza vaccination** |  |  |
| • Yes | 44 | 90.1 |
| • No | 400 | 9.9 |
| **Duration since symptoms onset, Days** |  |  |
| Mean± SD (Range) | 35.31±18.75 (10-120) |  |
| **Quarantine status** |  |  |
| • Hospital | 114 | 25.7 |
| • Home | 330 | 74.3 |
| **Oxygen supplementation** |  |  |
| • Yes | 92 | 20.7 |
| • No | 352 | 79.3 |
| **Intensive care unit admission** |  |  |
| • Yes | 60 | 13.5 |
| • No | 384 | 86.5 |

♦ **Cardiovascular disorders** chronic heart disease, atrial fibrillation, heart failure, stroke, hypertension), **endocrine disorders** ( diabetes mellitus, thyroid disease), kidney disorders , chronic obstructive pulmonary disease, active cancer , immune disorders

### Post COVID-19 Functional Status Scale (PCFS)

Most of participants (63.1%) had a trivial limitation in activities after recovery from COVID-19 (Grade 1), 14.1% had slight (Grade 2), 2.5% had moderate (Grade 3), and only (0.5%) had severe functional limitation (Grade 4). Only 20% had no functional limitations (Grade 0) as shown in ***Table (2)***.

**Table 2:** Post COVID-19 Functional Status Scale (PCFS) in the studied recovered COVID - 19 cases (n=444)

| Variable | Frequency<br>(n=444) | Percentage<br>% |
| --- | --- | --- |
| No limitation (Grade 0) | 89 | 20.0 |
| Negligible limitation (Grade 1) | 280 | 63.1 |
| Slight limitation (Grade 2) | 64 | 14.4 |
| Moderate limitation (Grade 3) | 9 | 2.0 |
| Severe (Grade 4) | 2 | 0.5 |
| Total | 444 | 100.0 |

Regarding the association between both demographic and clinical features of study group and their PCFS, there was a substantial variance between the grade of functional activity limitation (based on PCFS score) with age (P = 0.003), gender (P = 0.014), the duration since

COVID-19 symptoms onset (P <0.001), need for oxygen supplementation (P <0.001), ICU admission (P = 0.003), seasonal influenza vaccination (P <0.001), smoking status (P < 0.001) and lastly the presence of any comorbid disorders (P <0.001) ***Table (3)***.

**Table 3:** Association between Demographic and Clinical characteristics and POST COVID-19 Functional Status Scale (PCFS)in the studied recovered COVID-19 cases (n=444)

| Variable | Grade 0 |  | Grade 1 |  | Grade 2 |  | Grade 3 |  | Grade 4 |  |
| --- | --- | --- | --- | --- | --- | --- | --- | --- | --- | --- |
|  | N | % | N | % | N | % | N | % | N | % |
| **Age ( years)** |  |  |  |  |  |  |  |  |  |  |
| Mean ± SD | 30.06±10.28 |  | 33.11±11.73 |  | 36.62±14.12 |  | 37.33±18.35 |  | 32.50±6.36 |  |
| P value | **0.003<sup>#</sup>** |  |  |  |  |  |  |  |  |  |
| **Gender** |  |  |  |  |  |  |  |  |  |  |
| • Male | 50 | 26.0 | 120 | 62.5 | 19 | 9.9 | 2 | 1.0 | 1 | 0.5 |
| • Female | 39 | 15.5 | 160 | 63.5 | 45 | 17.9 | 7 | 2.8 | 1 | 0.4 |
| P value | **0.014<sup>##</sup>** |  |  |  |  |  |  |  |  |  |
| **Residence** |  |  |  |  |  |  |  |  |  |  |
| • Urban | 59 | 18.7 | 212 | 67.1 | 38 | 12.0 | 6 | 1.9 | 1 | 0.3 |
| • Rural | 30 | 23.4 | 68 | 53.1 | 26 | 20.3 | 3 | 2.3 | 1 | 0.8 |
| P value | **0.069** |  |  |  |  |  |  |  |  |  |
| **Duration since symptoms onset in days** |  |  |  |  |  |  |  |  |  |  |
| Mean ± SD | 38.87±17.69 |  | 34.52±19.01 |  | 33.67±17.79 |  | 38.89±26.00 |  | 25.00±14.14 |  |
| P value |  | < 0.001<sup>#</sup> |  |  |  |  |  |  |  |  |
| Quarantine status |  |  |  |  |  |  |  |  |  |  |
| • Hospital | 17 | 14.9 | 76 | 66.7 | 17 | 14.9 | 3 | 2.6 | 1 | 0.9 |
| • Home | 72 | 21.8 | 204 | 61.8 | 47 | 14.2 | 6 | 1.8 | 1 | 0.3 |
| P value |  | 0.516 |  |  |  |  |  |  |  |  |
| Oxygen supplementation |  |  |  |  |  |  |  |  |  |  |
| • Yes | 0 | 0.0 | 70 | 76.1 | 19 | 20.7 | 2 | 2.2 | 1 | 1.1 |
| • No | 89 | 25.3 | 210 | 59.7 | 45 | 12.8 | 7 | 2.0 | 1 | 0.3 |
| P value |  | < 0.001<sup>##</sup> |  |  |  |  |  |  |  |  |
| Intensive care unit admission |  |  |  |  |  |  |  |  |  |  |
| • Yes | 2 | 3.3 | 42 | 70.0 | 14 | 23.3 | 1 | 1.7 | 1 | 1.7 |
| • No | 87 | 22.7 | 238 | 62.0 | 50 | 13.0 | 8 | 2.1 | 1 | 0.3 |
| P value |  | 0.003<sup>##</sup> |  |  |  |  |  |  |  |  |
| Seasonal influenza vaccination |  |  |  |  |  |  |  |  |  |  |
| • Yes | 89 | 22.3 | 280 | 70.0 | 31 | 7.8 | 0 | 0.0 | 0 | 0.0 |
| • No | 0 | 0.0 | 0 | 0.0 | 33 | 75.0 | 9 | 20.5 | 2 | 4.5 |
| P value |  | < 0.001<sup>##</sup> |  |  |  |  |  |  |  |  |
| Smoking status |  |  |  |  |  |  |  |  |  |  |
| • None | 80 | 23.1 | 214 | 61.8 | 47 | 13.6 | 4 | 1.2 | 1 | 0.3 |
| • Active | 9 | 15.5 | 30 | 51.7 | 16 | 27.6 | 2 | 3.4 | 1 | 1.7 |
| • Former | 0 | 0.0 | 36 | 90.0 | 1 | 2.5 | 3 | 7.5 | 0 | 0.0 |
| P value |  | < 0.001<sup>##</sup> |  |  |  |  |  |  |  |  |
| Presence of any comorbidity |  |  |  |  |  |  |  |  |  |  |
| • Yes | 0 | 0.0 | 36 | 32.4 | 64 | 57.7 | 9 | 8.1 | 2 | 1.8 |
| • No | 89 | 26.7 | 244 | 73.3 | 0 | 0.0 | 0 | 0.0 | 0 | 0.0 |
| P value |  | < 0.001<sup>##</sup> |  |  |  |  |  |  |  |  |

<sup>#</sup>One-way ANOVA test
<sup>##</sup>Chi-square test

## Discussion

During the pandemic of COVID-19, we have been encountered with an enormous proportion of cases with diverse clinical features such as cough, fever, shortness of breath, musculoskeletal (lethargy & joint ache), gastro-intestinal, and sleep disorders **(13, 14, 15)**. However, evidence is missing on the functional state after recovery. As far as we know, this is the first report to assess the persistent restrictions of functional activity among convalescent COVID-19 cases using the recommended PCFS. We found that 80% of COVID-19 recovered cases have diverse degrees of functional restrictions ranging from negligible (63.1%), slight (14.4%), moderate (2%) to severe (0.5%) based on PCFS. Furthermore, there was a substantial variance between the score of PCFS with age (P= 0.003), gender (P= 0.014), the duration since the onset of the symptoms of COVID-19 (P <0.001), need for oxygen supplementation (P<0.001), ICU admission (P= 0.003), previous periodic influenza vaccination (P<0.001), smoking status (P < 0.001) and lastly the presence of any comorbid disorders (P <0.001).

These results are not surprising as, in addition to the impairment in physical activities, the long duration of confinement and the extreme doubt during the COVID-19 disease had generated remarkable mental and attitude disorders **(16)**.

In accordance with current results, several patients in the convalescence phase of SARS suffered from restrictions in physical activity causing fluctuating grades of restrictions in their work-related, public, and vacation activities or circadian living activities **(17, 18)**. It was found that the exercise capability and physical status of SARS recovered cases were considerably worse than that of normal publics after 6 month follow up. The functional frailty seemed disproportionate to the degree of functional lung injury and may be associated with additional aspects such as muscle weakness **(19)**. It was concluded that recovered cases of SARS, had outstanding defects identified with pulmonary function testing, with DLCO aberrations up to two years after retrieval, along with the health-related quality of life deficit **(6)**. Lastly, the long-standing hazardous properties of common enduring pain, lethargy, emotional stress, and troubled sleep after severe SARS led to the inability to return to dynamic effort for a minimum of one year after their acute disease **(20)**.

Similarly, the MERS convalescent cases also reported the ominously lower quality physical health for at least 14 months after infection start, also survivors who anticipated intensive care unit admittance described the ominously minor inclusive quality of life than those with non-critical disease **(21)**.

In the present study, only 3% of cases necessitating ICU did not record any functional restriction and 93.3% had negligible to slight functional restriction (compared to 22.7% no restrictions and 75% negligible-slight functional restriction, in patients not admitted in ICU, P= 0.003). It was recorded that patients who require intensive care admittance or even invasive mechanical ventilation are at great hazard for emerging post-intensive care syndrome (PICS) **(22)**. It is a usually detected phenomenon inside ICU recovered cases among the different age groups and often is described as protracted incapacity consequential to muscle dysfunction, lethargy, pain, and shortness of breath **(23)**.

It is recommended that the functional state could have predictive value for COVID-19 patients, as compromised physical activity was independently concomitant with worst consequences in hospitalized cases with community-acquired pneumonia, according to a recent prospective study **(24)**. The performance status may forecast one-month death rates as well as the frequently used CRB-65 score (confusion, respiratory rate, blood pressure, and age ≥ 65) in patients with any bacteriological or viral pneumonia **(25)**. So, the incorporation of patients’ functional status measurement into patient assessment may improve the prognostic ability of current risk classification systems to predict mortality from COVID-19 pneumonia **(26)**. The use of simple scales as suggested by Klok and colleagues **(12)** may be very important in the assessment and follow-up of functional status in this novel Post-COVID-19 syndrome and may reduce its related morbidities.

Since March, 2020 reports indicated that the severity and outcome of COVID-19 pneumonia (SARS-CoV-2) is affected by patients age, gender, smoking, ICU admission, previous comorbidities **(1,13,14,26,27)**. To the best of our knowledge this is the first report of persistent effect of these factors on the functional status of COVID-19 after recovery. The exact mechanism is not yet explained.

**Limitations of the study:** first, the lack of data of functional status before COVID-19 infection; second, the history of the symptoms both at the onset of COVID-19 and after recovery is not included; third, the pharmacologic therapy given to the patients was not mentioned (however all patients received the standard protocol of Ministry of health and population in Egypt but it was changed several times according to international recommendations), lastly, random selection bias may be present and an inability for personal face-to-face interview in some cases.

### Conclusions

Most of COVID-19 recovered cases have different degrees of functional limitations ranging from negligible to severe based on PCFS. These limitations were affected by age, gender, periodic influenza vaccination, smoking status, duration since symptoms onset, need for oxygen therapy or ICU admittance, and lastly the presence of coexisting comorbidity.

It is recommended that Post COVID-19 monitoring programs should be implemented in specific clinical settings or as an out-patients program to follow the functional status of patients in 1, 3, 6 months visits to support the complete care for cases recovered from COVID-19. Furthermore, extended monitoring using simple scales as PCFS is necessary to determine whether these functional deficits after COVID-19 recovery persist or not. Further studies are required to explain the underlying cause of post COVID-19 functional limitation.

## Data Availability

on request

